# Interpretable Machine Learning for Predicting Multiple Sclerosis Conversion from Clinically Isolated Syndrome

**DOI:** 10.1101/2024.07.18.24310578

**Authors:** Eden Caroline Daniel, Santosh Tirunagari, Karan Batth, David Windridge, Yashaswini Balla

**Affiliations:** Department of Computer Science, Middlesex University, The Burroughs, London NW4 4BT; West Suffolk Hospital NHS Foundation Trust, Hardwick Ln, Bury Saint Edmunds IP33 2QZ; Princess Royal Hospital, SaTH NHS Trust, Apley Castle, Telford TF1 6TF

**Keywords:** Clinically isolated syndrome, Multiple sclerosis, Machine learning, Predictive modelling, CIS to MS conversion

## Abstract

**Background:** Machine learning (ML) prediction of clinically isolated syndrome (CIS) conversion to multiple sclerosis (MS) could be used as a remote, preliminary tool by clinicians to identify high-risk patients that would benefit from early treatment.

**Objective:** This study evaluates ML models to predict CIS to MS conversion and identifies key predictors.

**Methods:** Five supervised learning techniques (Naïve Bayes, Logistic Regression, Decision Trees, Random Forests and Support Vector Machines) were applied to clinical data from 138 Lithuanian and 273 Mexican CIS patients. Seven different feature combinations were evaluated to determine the most effective models and predictors.

**Results:** Key predictors common to both datasets included sex, presence of oligoclonal bands in CSF, MRI spinal lesions, abnormal visual evoked potentials and brainstem auditory evoked potentials. The Lithuanian dataset confirmed predictors identified by previous clinical research, while the Mexican dataset partially validated them. The highest F1 score of 1.0 was achieved using Random Forests on all features for the Mexican dataset and Logistic Regression with SMOTE Upsampling on all features for the Lithuanian dataset.

**Conclusion:** Applying the identified high-performing ML models to the CIS patient datasets shows potential in assisting clinicians to identify high-risk patients.

## Introduction

Multiple Sclerosis (MS) is an autoinflammatory, demyelinating disorder of the central nervous system (CNS), affecting 165 people per 100,000 in North America (1) and 2.8 million worldwide (2). A precursory stage called Clinically Isolated Syndrome (CIS) often precedes Clinically Definite Multiple Sclerosis (CDMS). In CIS, patients experience an episode of neurological symptoms, with features of MS affecting the optic nerves, brain or spinal cord, for at least 24 hours (3, 4).

Not all MS patients have a CIS stage, although the vast majority (85%) do, whereas 37% of CIS patients do not convert to MS even after 20 years (5, 6).

Diagnosis is *disseminated in time and space*, referring to evidence of damage to different CNS loci and the disease’s relapsing nature over a patient’s life course. MS aetiology is complex involving gene-environment interactions and risk factors including: female gender, Epstein-Barr virus exposure, smoking, low vitamin D level and childhood obesity (7). Protective factors include: breastfeeding for more than four months, longer schooling years and socioeconomic status (8). Risk factors are not routinely accounted for in diagnostic workflows.

Currently, diagnosis is made manually using Macdonald 2017 criteria (9) taking into account clinical features (from examination and investigation), biomarkers (such as oligoclonal bands in cerebrospinal fluid indicating CNS damage) and magnetic resonance imaging (MRI) lesions typical of MS (9, 10). This is a subjective, user-dependent process that can introduce inconsistencies (11). CIS to MS conversion, which may take months to years (12), is a critical entry-point for disease-modifying treatments in high-risk individuals to delay onset and reduce disability (13, 14). Predicting highrisk vs low-risk individuals allows for individualised treatment and has high clinical utility (15).

Machine Learning (ML) represents a rapid and inexpensive alternative to manual diagnosis in a number of medical settings (16). Presently, while ML techniques have the potential to disentangle the complex relationship between CIS and MS they require proofs of reliability and interpretability prior to clinical implementation (17, 18). Studies predicting conversion using ML have primarily relied on MRI scans. Zhang et al. (19) used random forests (RF) classifiers on lesion shape features achieving 84.5% accuracy, exceeding the 75% baseline for the original McDonald (2010) criteria. The study was limited by database size and lack of multimodal data. Yoo et al. (20) combined CNN-extracted lesion features with clinical data for 75% accuracy but faced challenges with small sample size and incomplete follow-up. Probert et al. (6) employed a multi-omics algorithm on CSF biomarkers achieving 83% accuracy but did not account for MRI lesions and other non-CSF features. Wottschel et al. (21) combined lesion features, demographics, and clinical information using Support Vector Machines (SVM) model which achieved 71.4% accuracy.

Recently, Rasouli and colleagues (22) sought to apply an explainable artificial intelligence/ML (XAI) (23) approach to a publically-available Mexican CIS dataset and achieved accuracy between 78.3% and 83.6% (24). An additional, retrospective, Lithuanian CIS dataset (25) is also publically available and we make use of this. These Mexican and Lithuanian studies appraised predictors manually and demonstrated CIS to MS conversion rates of 46% within 10 years and 35.5% within 5 years respectively.

In the following study, we will build on this work by making use of routinely collected data (clinical, demographic and MRI), presenting five machine learning classification algorithms for prediction of CIS to MS conversion. The models, chosen for their simplicity to hone in on appraising feature combinations, are: *Naïve Bayes* (NB, serving as a lower benchmark), *logistic regression* (LR), *decision trees* (DT), *Random Forest* (RF) and *Support Vector Machine* (SVM). They are ordered in their observed performance on arbitrary, small data problems.

By exploring multiple algorithms we aim to reduce the inductive bias arising from use of a single algorithm. The ML algorithms will be applied to the two indicated datasets and also to their concatenation in order to establish generalizability.

### Objectives

The objectives of this study are as follows:

- **CIS to MS conversion prediction using ML** with various feature combinations on two different datasets and on their concatenation.
- **Understanding interpretability** by determining the model-feature combination with the highest F1 score.
- **Comparing ML-identified predictors** with original study predictors as well as prior research.
- **Validating clinical usefulness** by assessing generalizability.

### Scope and Limitations

Publicly-available datasets are chosen, over hospital data collection, for their diversity of features, suitability for the objectives, and capacity for replicability. Deep learning (DL) is not used due to the small dataset size and its tendency to obscure feature relevance. The study is inherently constrained by its sample size (total patients = 411) and the many missing values in the Lithuanian dataset.

As the datasets are from two different populations, common conversion predictors independent of demographic influence can be identified to enhance the generalizability and clinical usefulness of the identified ML models and features. However, the population-specific nature of each dataset may introduce selection bias.

To mitigate small sample size issues, imputation, SMOTE (synthetic minority oversampling technique) Upsampling, dataset concatenation are employed. A larger dataset with at least 1000 observations would improve model training.

## Materials and Methods

### Datasets

Two publicly available datasets were utilized, both downloadable on Mendeley Data via the *CC BY 4*.*0 license*. The ethical approval for using the datasets is available in the Supplementary Material: Fig. S11.

The first dataset^1^ was taken from a prospective study (24) comprising 273 Mexican CIS patients with 20 features who presented to the National Institute of Neurology and Neuroscience, Mexico, between 2006 and 2010. The second dataset^2^ was sourced from a retrospective study (25) comprising 138 Lithuanian CIS patients with 44 features who presented to the Hospital of Lithuanian University of Health Sciences, Lithuania, between 2015 and 2020.

### Analysis of datasets

The datasets contain demographic, clinical, and MRI features in binary format. Features common to both datasets are: age, gender, visual evoked potentials positive (VEP+), brainstem auditory evoked potentials positive (BAEP+), oligoclonal bands positive (OCB+) in CSF, MRI spinal, infratentorial and periventricular lesions and MS outcome (refer to Fig. 1). Exclusive features in the Mexican dataset include: initial and final expanded disability status scale EDSS, breastfeeding status, schooling years, upper limb and lower limb somatosensory evoked potentials (ULSSEP and LLSSEP, respectively), varicella zoster infection and cortical lesions on MRI. Exclusive features in the Lithuanian dataset include: proprioception, coordination, muscle strength, Rossolimo and Babinski’s signs, vertigo, urinary retention, immunoglobulin levels in CSF and juxtacortical lesions on MRI. Further detail comparing features between the datasets can be found in the Supplementary Material section (Data Analysis).

**Fig. 1.**
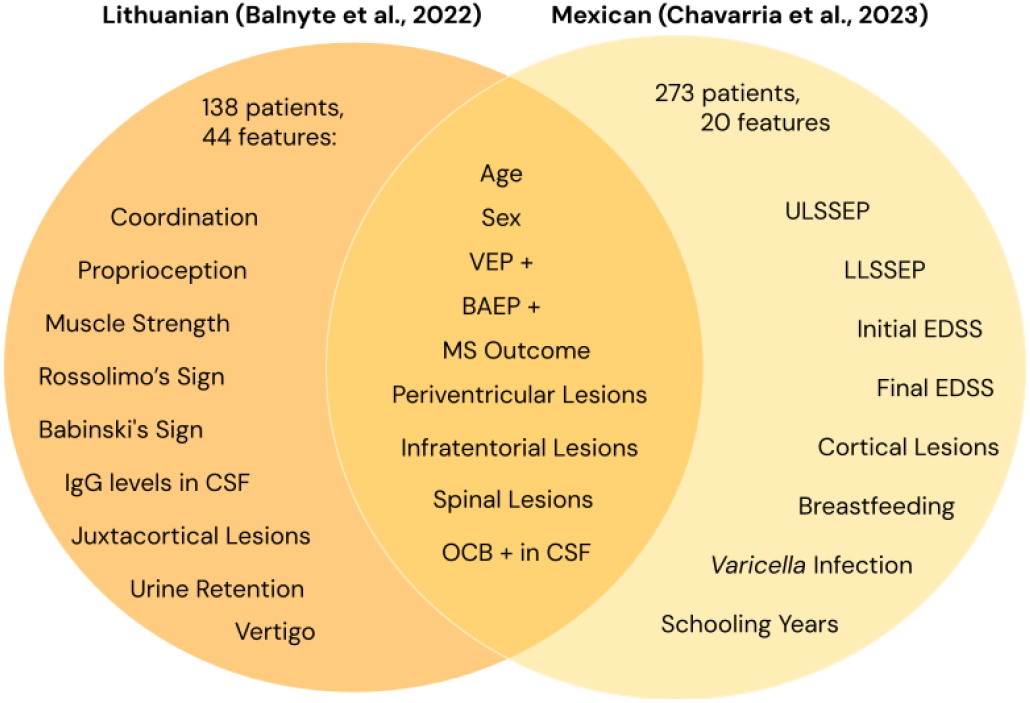
Venn diagram showing the features in both datasets and common features.

### Data preprocessing

Data cleaning involved replacing empty cells with ‘not-a-number’ values to ensure data integrity. All columns with categorical values were transformed to a numerical format for consistency. Data was structured for model use, focusing on the target variable representing the presence or absence of multiple sclerosis. When SMOTE upsampling and principle component analysis (PCA) were applied, they generated resampled and PCA transformed data. The data was then divided into 80% training and 20% test sets, allowing the model to learn from a substantial portion (training set) and evaluate performance on unseen data (test set).

### ML classifiers

Classification in machine learning can be binary or multiclass (23). The classifiers chosen for this study are simple and interpretable. Naive Bayes (NB) is a probabilistic algorithm exploiting Bayes’ theorem under an assumption of feature independence. It is valued for its simplicity and efficiency, and generally deployed as a lower bound on performance (26). Logistic Regression (LR) uses logistic functions to predict class probabilities, offering interpretability and stochastic robustness but is limited by its linear assumption (27). Decision Trees (DTs) provide high interpretability and resilience to outliers but may overfit and struggle with complex data (28). Random Forests (RFs) improve upon DTs by aggregating multiple trees through attribute bootstrap aggregation, handling high-dimensional data well but requiring additional hyperparameteric tuning (29). Support Vector Machines (SVMs) obtain maximum-margin decision boundaries, excelling in high-dimensional spaces but are computationally intensive and less interpretable. This makes them a presumed *a priori* upper benchmark (16, 30). They are also readily extended in functionality via *kernelization* (though beyond the scope of this work).

### Structural Risk Management

These methods reduce overfitting, model complexity, and improve generalization to unseen data.

Class imbalances were handled using stratification during the 80:20 data split to maintain proportional class distribution in both sets. SMOTE Upsampling was applied to the best model-feature combination, creating synthetic samples for the minority class by interpolating between existing samples. This balanced the class distribution, preventing bias towards the majority class and improving performance on the underrepresented class. In the Lithuanian dataset, the majority to minority ratio was 64.5% No MS to 35.5% MS. In the Mexican dataset, it was 54.4% No MS to 45.6% MS. Applying SMOTE Upsampling balanced the classes to a 50:50 distribution.

To handle missing values, three imputation techniques were applied: Simple Imputation (SI), Expectation-Maximisation (EM), and Multiple Imputation by Chained Equations (MICE). These ensure data integrity and minimize bias without reducing dataset size.

Stratified K-fold Cross-validation (K=5) evaluated model performance and generalization, balancing class distribution in each fold. Hyperparameter tuning was conducted using GridSearchCV optimizing parameters for all models to enhance performance. The best estimator was then trained on the training set and used to make predictions.

Evaluation utilizes F1 score, a standard classification metric, to assess model performance. The best value of F1 score is 1 and the worst is 0. Features Importance Scores identified influential predictors in the models.

### Experiments and Results

The overall pipeline involved data loading, preprocessing, structural risk management, model training, prediction, evaluation and identification of conversion predictors. The models used included NB as a lower benchmark, LR and DT for simplicity and interpretability, RF to enhance DT performance, and SVM as an upper benchmark. The experiments tested various imputation methods, PCA and feature selection to determine the optimal model-feature combination. Finally, SMOTE Upsampling and cross-dataset validation were performed to assess model performance and generalizability (hence indicating clinical usefulness).

#### 1. Effect of Imputation

Missing values in datasets have the potential to cause integrity issues and hamper overall model accuracy. To address this, we employ industry-standard imputation methods to fill in missing values prior to model fitting: SI, EM and MICE (as presented in Supplementary Material: Table S1.) These were tested on the Lithuanian dataset for NB, DT, and RF models. Using SI, DT achieved the highest test F1 score compared to the rest. The EM Imputer yielded similar test F1 scores across all models. MICE exhibited overfitting with respect to RF but retained consistent F1 scores for NB and DT. Overall, EM was the superior imputation method and DT was the best-performing model across all imputations by being most consistent.

#### 2. Effect of Multicollinearity

Multicollinearity among features can negatively impact model performance by inflating variances and causing models to be unstable. To address this, we aim to determine whether the application of PCA, which mitigates multicollinearity by determining principal components of variation, can improve model performance. Models were evaluated with and without PCA to assess its impact. The results are presented in Supplementary Material: Table S2. PCA produced overfitting in all models across all imputations, with MICE giving slightly better results. We conclude that the application of PCA deteriorates model performance in this data context, with little evidence of multicolinearity or stochastic noise in the features.

#### 3. Effect of Automatic Feature Selection

Automatic feature selection can streamline the modeling process by identifying and using only the most relevant features. To test this, we evaluate whether automatic feature selection during model training improves model performance as compared to the use of all features without automatic selection. As shown in Table 1, SI automatic feature selection improved F1 scores but did not alleviate overfitting (other than for RF without PCA). For EM, it improved F1 test scores, both with and without PCA, but overfitted in all cases. For MICE, it produced no change in DT F1 scores but reduced test overfitting with respect to RF. Overall, automatic feature selection did not mitigate overfitting. Automatic feature selection on MICE and EM imputations gave similar model performances.

**Table 1.**
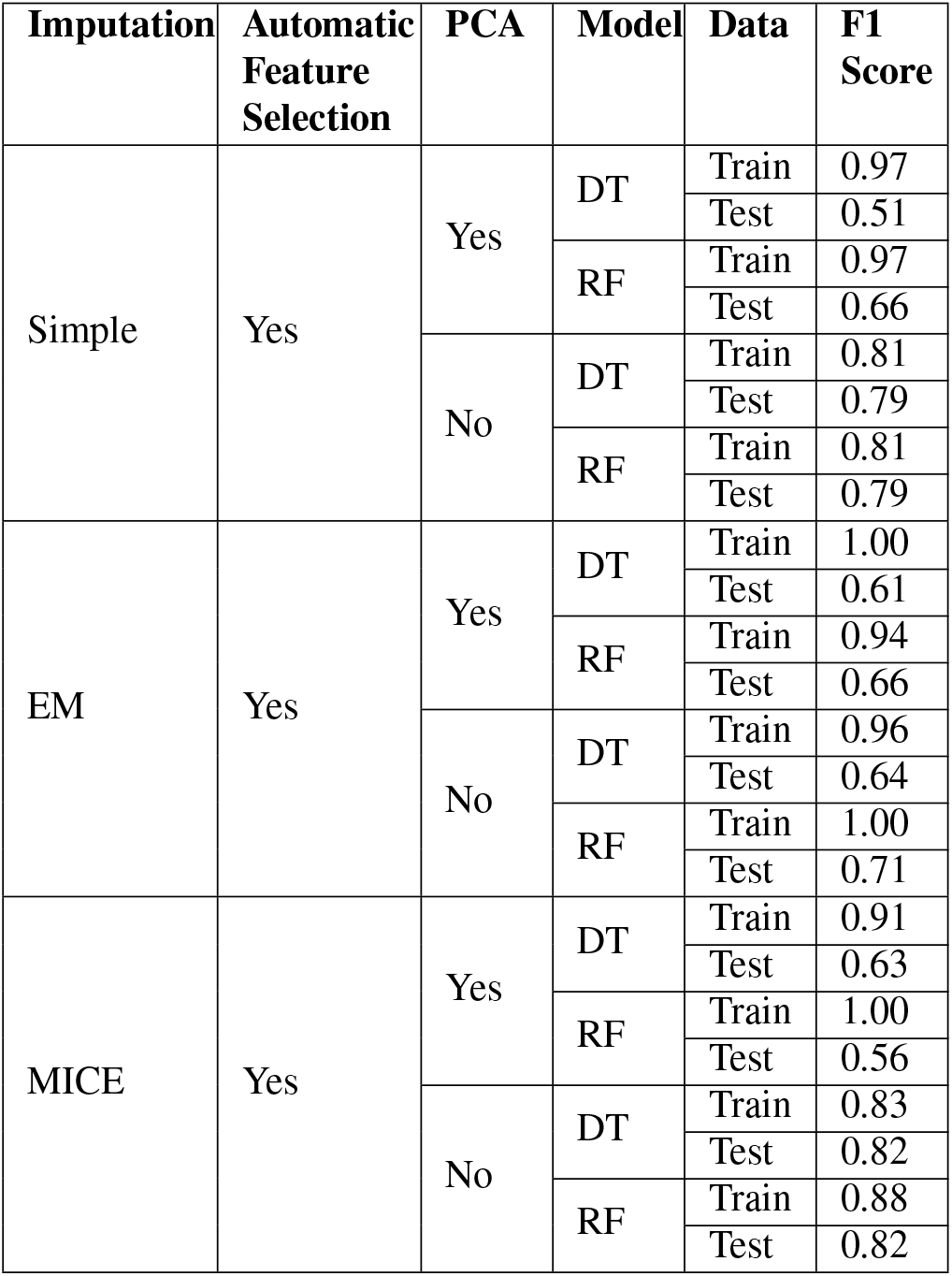
Effect of automatic feature selection on model F1 Scores across various imputations, with and without PCA.

| Imputation | Automatic Feature Selection | PCA | Model | Data | F1 Score |
| --- | --- | --- | --- | --- | --- |
| Simple | Yes | Yes | DT | Train | 0.97 |
|  |  |  |  | Test | 0.51 |
|  |  |  | RF | Train | 0.97 |
|  |  |  |  | Test | 0.66 |
|  |  | No | DT | Train | 0.81 |
|  |  |  |  | Test | 0.79 |
|  |  |  | RF | Train | 0.81 |
|  |  |  |  | Test | 0.79 |
| EM | Yes | Yes | DT | Train | 1.00 |
|  |  |  |  | Test | 0.61 |
|  |  |  | RF | Train | 0.94 |
|  |  |  |  | Test | 0.66 |
|  |  | No | DT | Train | 0.96 |
|  |  |  |  | Test | 0.64 |
|  |  |  | RF | Train | 1.00 |
|  |  |  |  | Test | 0.71 |
| MICE | Yes | Yes | DT | Train | 0.91 |
|  |  |  |  | Test | 0.63 |
|  |  |  | RF | Train | 1.00 |
|  |  |  |  | Test | 0.56 |
|  |  | No | DT | Train | 0.83 |
|  |  |  |  | Test | 0.82 |
|  |  |  | RF | Train | 0.88 |
|  |  |  |  | Test | 0.82 |

#### 4. Manual Feature Selection

Identifying the most relevant features manually can improve interpretability. To achieve this, we aimed to determine which feature combinations maximize model performance. Various combinations as presented including original study features, all features and features except multicollinear ones were tested. In the Lithuanian dataset (see Table 2), the best test model-feature combination was LR using all features, with an F1 score reaching 0.89. For the Mexican dataset, the best model was RF using all features with a test score of 1.00 (see Table. 3).

**Table 2.** F1 scores of all five models across various feature combinations for the Lithuanian dataset.

| Features | Data | NB | LR | DT | RF | SVM |
| --- | --- | --- | --- | --- | --- | --- |
| All | Train | 0.59 ± 0.08 | 0.97 ± 0.03 | 0.73 ± 0.12 | 0.93 ± 0.05 | 0.94 ± 0.07 |
|  | Test | 0.59 | 0.89 | 0.51 | 0.82 | 0.79 |
| All Except Multicollinear | Train | 0.67 ± 0.07 | 0.77 ± 0.06 | 0.73 ± 0.10 | 0.95 ± 0.06 | 0.80 ± 0.06 |
|  | Test | 0.59 | 0.78 | 0.82 | 0.71 | 0.68 |
| Original Study | Train | 0.73 ± 0.04 | 0.75 ± 0.03 | 0.74 ± 0.03 | 0.78 ± 0.05 | 0.76 ± 0.04 |
|  | Test | 0.82 | 0.82 | 0.86 | 0.82 | 0.86 |

**Table 3.** F1 scores of all five models across various feature combinations for the Mexican dataset.

| Features | Data | NB | LR | DT | RF | SVM |
| --- | --- | --- | --- | --- | --- | --- |
| All | Train | 0.73 ± 0.02 | 0.84 ± 0.02 | 0.96 ± 0.01 | 0.98 ± 0.01 | 0.83 ± 0.03 |
|  | Test | 0.80 | 0.87 | 0.98 | 1.00 | 0.85 |
| All Except Multicollinear | Train | 0.73 ± 0.02 | 0.85 ± 0.02 | 0.75 ± 0.04 | 0.94 ± 0.02 | 0.87 ± 0.07 |
|  | Test | 0.80 | 0.87 | 0.84 | 0.87 | 0.85 |
| Original Study | Train | 0.60 ± 0.03 | 0.59 ± 0.03 | 0.44 ± 0.23 | 0.60 ± 0.02 | 0.60 ± 0.03 |
|  | Test | 0.64 | 0.64 | 0.39 | 0.64 | 0.71 |

#### 5. Effect of Upsampling

Class imbalance can bias models towards the majority class, reducing performance on the minority class. To address this, we aimed to improve training accuracy by enhancing minority class representation. SMOTE Upsampling was applied to the best-performing models to balance the class distribution and improve model performance. In the Lithuanian dataset, SMOTE applied to the best model (LR using all features) gave perfect F1 scores of 1.00 for both train and test sets (Table 4). In the Mexican dataset, SMOTE on the best model (RF using all features) increased train scores to 0.99 but decreased test scores to 0.95 (Table 4).

**Table 4.** Effect of SMOTE Upsampling on best model F1 scores in both datasets.

| Dataset | Feature Combination | Data | Scores |
| --- | --- | --- | --- |
| Lithuanian | LR (ALL) | Train | 1.00 ± 0.00 |
|  |  | Test | **1.00** |
| Mexican | RF (ALL) | Train | 0.99 ± 0.01 |
|  |  | Test | **0.95** |

#### 6. Cross-Dataset Validation

Evaluating model generalizability to new populations helps detect biases and improve robustness. This is integral for clinical application. Models were trained on the Lithuanian dataset and tested on the Mexican dataset, and vice versa. There was also further validation on their concatenated dataset (i.e. combining observations from both).

Training on the Lithuanian dataset and cross-validating on the Mexican caused severe overfitting with a test F1 score of 0.56, indicating poor generalizability. Training on the Mexican and cross-validating on the Lithuanian gave better results with a test F1 score of 0.82 and good generalization to unseen data. The best model for training on Lithuanian and testing on Mexican, as well as for training on Mexican and testing on Lithuanian was LR (Table 5).The best model on the concatenated dataset was LR with a test F1 score of 0.72 (Table 6).

**Table 5.** F1 Scores for cross-dataset validation.

| Dataset | Data | NB | LR | DT | RF | SVM |
| --- | --- | --- | --- | --- | --- | --- |
| Train on Lithuanian, Test on Mexican | Train | 0.76 ± 0.03 | 0.80 ± 0.03 | 0.89 ± 0.06 | 0.93 ± 0.05 | 0.82 ± 0.05 |
|  | Test | 0.55 | 0.56 | 0.40 | 0.41 | 0.45 |
| Train on Mexican, Test on Lithuanian | Train | 0.61 ± 0.02 | 0.64 ± 0.05 | 0.62 ± 0.04 | 0.66 ± 0.04 | 0.63 ± 0.02 |
|  | Test | 0.78 | 0.82 | 0.76 | 0.81 | 0.63 |

**Table 6.** F1 Scores for concatenated dataset validation.

| Dataset | Data | NB | LR | DT | RF | SVM |
| --- | --- | --- | --- | --- | --- | --- |
| Concatenation | Train | 0.62 ± 0.01 | 0.64 ± 0.02 | 0.65 ± 0.04 | 0.67 ± 0.03 | 0.63 ± 0.02 |
|  | Test | 0.70 | 0.72 | 0.56 | 0.71 | 0.71 |

## Discussion

This study automated CIS to MS conversion prediction. We used five supervised machine learning algorithms on clinical, demographic, and MRI data derived from two CIS databases. The results are appraised for their accuracy (by way of F1 scores), as well as overfitting and generalisability (when there is sound accuracy on training data that reduces when applied to unseen, test data). We comment on the clinical implications of the experiments.

Several experiments were conducted to assess the impact of imputation methods on results. Simple averaging of missing values in SI led to overfitting in NB and RF, while EM improved performance for all models by capturing inter-feature relationships. The iterative approach of MICE maintained prediction consistency for NB and DT but caused RF to overfit. DT showed consistent performance across imputations. EM performed best without PCA or feature selection.

PCA was evaluated for its handling of dimensionality and multicollinearity among the 44 features in the Lithuanian dataset. However, it caused LR, DT, and RF models to overfit and perform poorly across all imputation methods. This was likely due to data loss, increased noise and the application of linear assumptions to non-linear data. Interestingly, PCA using MICE for imputation led to better F1 scores and reduced overfitting. Overall, however, PCA compromised interpretability, which is important for clinical use, and was thus not used in subsequent experiments.

Automatic feature selection aimed to reduce overfitting by identifying relevant features for predictive accuracy but did not consistently succeed across different imputations. It struggled with complex feature interactions, high data variability and noisy features. Although it enhanced F1 scores, it did not effectively combat overfitting so could not be used further.

Manual selection showed that all features worked best, as opposed to selecting the features used in the original studies (24, 25) or selecting all features excluding multicollinear ones. Regarding the best model-feature combination, for the Lithuanian dataset the best model was LR using all features. Following this was RF using all features. Feature importance scores (Fig. 2) revealed MRI spinal lesions, MRI lesions indicative of other diseases, general weakness and periventricular lesions as significant positive predictors for conversion from CIS to MS. Conversely, increased protein, Rossolimo sign and MRI lesions specific to MS were significant negative predictors. For the Mexican dataset, the best model was RF using all features (F1 test score 1.0), followed by DT using all features. Feature Importance Scores (Fig. 3) revealed periventricular lesions, initial symptoms and age as significant predictors for conversion from CIS to MS. Other important contributors included MRI infratentorial lesions, OCB+ in CSF and schooling. Using all features yielded the best results for most models across both datasets, most likely by capturing intricate feature interactions.

**Fig. 2.**
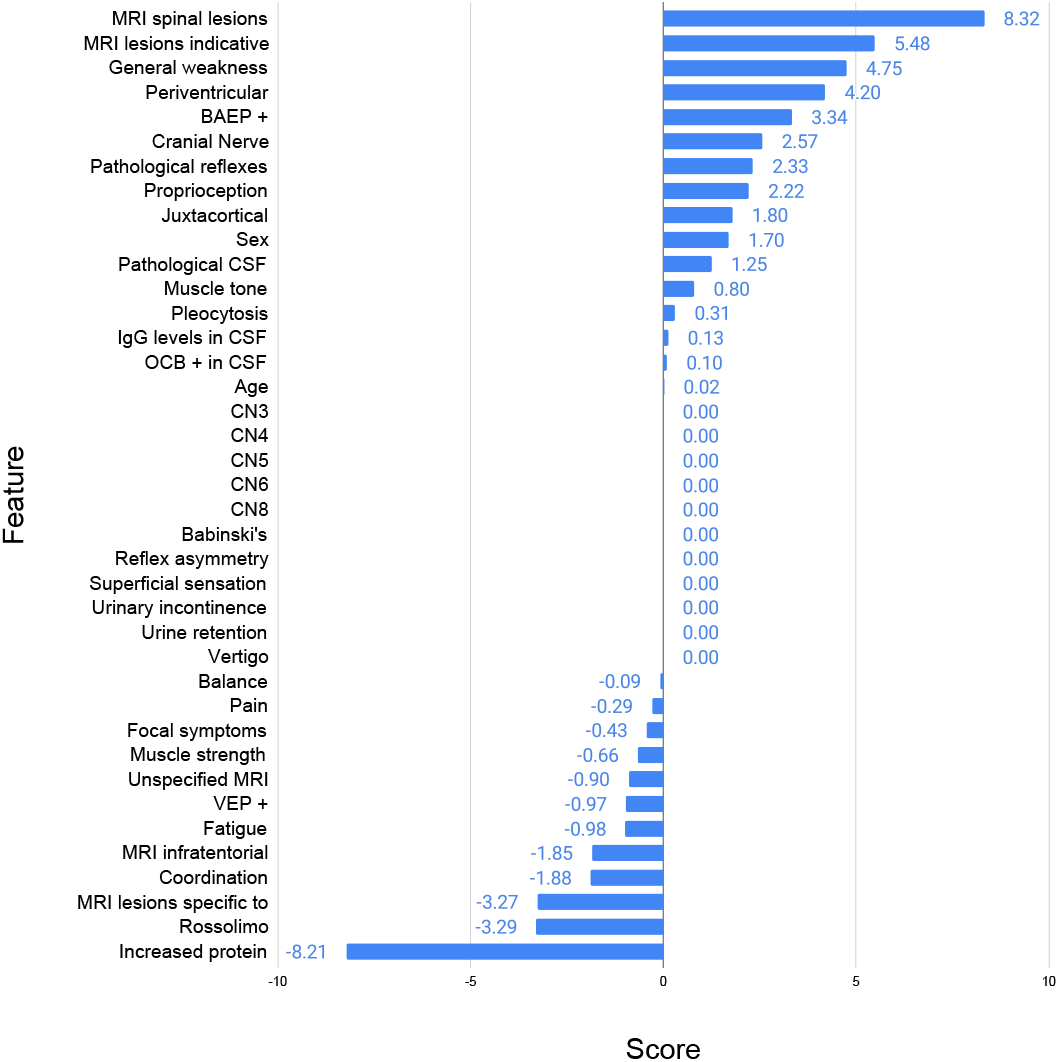
Feature Importance Scores for the best model, LR using all features on the Lithuanian dataset

**Fig. 3.**
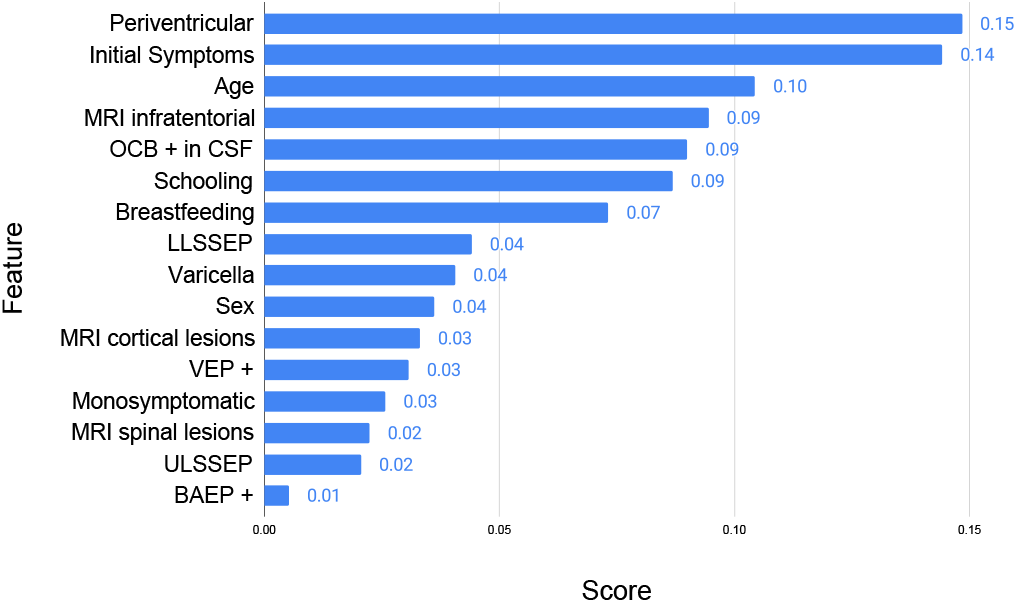
Feature Importance Scores for the best model, RF using all features on the Mexican dataset.

SMOTE upsampling, used to address small dataset size and class imbalance, exhibited divergent effects on the best models in both datasets. On the Lithuanian dataset, SMOTE increased the test F1 score of the best model (LR using all features) to 1.0, confirming its effectiveness. This result outperforms previous studies that cite accuracies ranging from 75% to 84.5% (6, 19, 20, 22). Conversely, on the Mexican dataset, SMOTE reduced the test F1 score of the best model (RF using all features) to 0.95 and overfitted. Overall, SMOTE achieved high accuracies and showed it may be useful in combatting bias towards majority classes.

When training on the Lithuanian dataset and cross-validating on the Mexican dataset, LR performed best. However, there was overfitting and hence poor generalisability (F1 score 0.56). LR also performed best when training on the Mexican dataset and cross-validating on the Lithuanian dataset (F1 score 0.82). This was superior to cross-validating the other way round. Even on the concatenated dataset, LR performed the best overall (F1 score 0.72). Differences in patient demographics, fewer observations in the Lithuanian dataset, and varying disease characteristics contributed to these discrepancies. The cross validation and concatenation experiments show appropriateness of LR in clinical contexts to predict CIS to MS conversion due to its generalizability.

The results show significant difference in the predictive abilities of all five models. For example, in the Lithuanian dataset using all features, the best model LR achieved a 0.89 test score, while the worst model NB performed at near chance levels. Similarly, in the Mexican dataset using original study features, SVM (the upper benchmark) scored 0.71, whereas DT performed the worst with a 0.39 test score. This emphasizes the significant impact of model choice on prediction accuracy and why trialling different models is essential in understanding the applicability of ML to clinical data.

This study validated the use of clinical features. This is evidenced by confirmation of predictors from previous studies that include OCB+ in CSF, BAEP+, MRI spinal lesions and proprioception (5, 31), as well as periventricular lesions, MRI spinal lesions, MRI infratentorial lesions, MRI cortical lesions and LLSSEP (32).

Experiments on the Lithuanian dataset confirmed all predictors manually identified in the original study (25). Additionally, MRI lesions indicative of other diseases, muscle tone abnormalities, sex, and VEP+ were identified as predictors. Conversely, the original Mexican study manually identified specific clinical parameters and MRI findings as predictors (24). However, the experiments on the Mexican dataset in this study validated different predictors, such as periventricular lesions, MRI spinal lesions, MRI infratentorial lesions, MRI cortical lesions and LLSSEP. We discovered predictors common to both datasets were sex, MRI spinal lesions, OCB+ in CSF, VEP+, and BAEP+, indicating their reliability as features. By confirming known predictors, for example MRI lesions and OCBs, that are used routinely by clinicians when applying the Macdonald criteria (9), we confirm holistic datasets that make use of multi-modal data improve reliability of ML methods; this in turn provides interpretability as clinicians would be able to understand the role of various features in these ML models.

This study also identified sex as a predictor, aligning with previous research (33, 34) but differing from Balnyt ė et al. (25). It found males more susceptible to MS conversion, contrary to previous research indicating higher female susceptibility. This may exemplify how small datasets can skew results and cautions the requirement for further study. Initial and final EDSS were identified as predictors, unlike as found by Chavarria et al. (24), which shows utility of wider data sources.

## Conclusion

The study aimed to evaluate ML methods in the prediction of CIS to MS conversion. Principal findings showed that EM and MICE imputations gave similar results, PCA deteriorated model performance and automatic feature selection did not mitigate overfitting.

The best models were LR using all features with SMOTE upsampling on the Lithuanian dataset and RF using all features on the Mexican dataset. Both achieved perfect F1 scores of 1.0. LR performed best during cross-validation and validation on concatenation. SMOTE Upsampling was useful for the best model on Lithuanian dataset but decreased the test score for the best model on Mexican dataset.

Common predictors for both datasets were OCB+ in CSF, sex, MRI spinal lesions, VEP+ and BAEP+. Predictors exclusive to the Mexican dataset were periventricular lesions, initial and final EDSS, MRI infratentorial lesions, MRI cortical lesions and LLSSEP. Exclusive predictors for the Lithuanian dataset were proprioception, MRI lesions indicative of other diseases and muscle tone abnormalities. The experiments on the Lithuanian dataset confirmed all predictors identified in the study by Balnytė et al. (25). However, the experiments on the Mexican dataset partially confirmed those identified manually by Chavarria et al. (24).

The models demonstrated clinical usefulness, with reasonable generalization when trained on the Mexican dataset. Despite discrepancies in identifying lesser-known predictors such as breastfeeding, schooling years and Varicella infection, this study validated widely-recognized predictors used by clinicians in MS diagnosis.

Future studies should consider larger, racially diverse datasets with at least 1000 observations. They should include risk factors such as Vitamin D levels, smoking, EpsteinBarr virus infection and teenage obesity, as well as genetic data (33). Deep learning models could be explored in larger datasets. Integrating the best-performing models into clinical practice may enhance CIS to MS conversion prediction. This would aid clinical decision-making to individualise treatment for CIS individuals who are at high-risk of converting to MS.

## Data Availability

Data availability. The datasets used in this study are available from Mendeley via the CC BY 4.0 License.
The Mexican dataset is available at https://data.mendeley.com/datasets/8wk5hjx7x2/1
and the Lithuanian dataset is available at https://data.mendeley.com/datasets/yjfydt34rs/1.
Additionally, we made the data available at https://github.com/tsantosh7/Multiple-Sclerosis-Conversion in the "Data" folder, which includes both datasets as well as the concatenated dataset.

https://data.mendeley.com/datasets/yjfydt34rs/1

https://github.com/tsantosh7/Multiple-Sclerosis-Conversion

## Availability and Implementation

### Code availability

The code for the machine learning algorithms and data analysis is available in the GitHub repository: https://github.com/tsantosh7/Multiple-Sclerosis-Conversion

### Data availability

The datasets used in this study are available from Mendeley via the CC BY 4.0 License. The Mexican dataset is available at https://data.mendeley.com/datasets/8wk5hjx7x2/1 and the Lithuanian dataset is available at https://data.mendeley.com/datasets/yjfydt34rs/1. Additionally, we made the data available at https://github.com/tsantosh7/Multiple-Sclerosis-Conversion in the “Data” folder, which includes both datasets as well as the concatenated dataset.

## Author contributions

E.C.D: Implementation, Analysis, Investigation, Visualization and Writing manuscript; S.T: Conceptualization, Methodology, Investigation, Supervsion; K.B: Writing manuscript, Proof Reading, Clinical Validation; D.W: Methodology, Supervsion; Y. B: Conceptualization, Writing Manuscript, Clinical Validation.

## Footnotes

1 https://data.mendeley.com/datasets/8wk5hjx7x2/1

2 https://data.mendeley.com/datasets/yjfydt34rs/1

## Bibliography

1. Mitchell T Wallin, William J Culpepper, Emma Nichols, Zulfiqar A Bhutta, Tsegaye Tewelde Gebrehiwot, Simon I Hay, Ibrahim A Khalil, Kristopher J Krohn, Xiaofeng Liang, Mohsen Naghavi, et al. Global, regional, and national burden of multiple sclerosis 1990–2016: a systematic analysis for the global burden of disease study 2016. The Lancet Neurology, 18 (3):269–285, 2019.

2. Clare Walton, Rachel King, Lindsay Rechtman, Wendy Kaye, Emmanuelle Leray, Ruth Ann Marrie, Neil Robertson, Nicholas La Rocca, Bernard Uitdehaag, Ingrid van Der Mei, et al. Rising prevalence of multiple sclerosis worldwide: Insights from the atlas of ms. Multiple Sclerosis Journal, 26(14):1816–1821, 2020.

3. David H Miller, Declan T Chard, and Olga Ciccarelli. Clinically isolated syndromes. The Lancet Neurology, 11(2):157–169, 2012.

4. Wallace J Brownlee and David H Miller. Clinically isolated syndromes and the relationship to multiple sclerosis. Journal of clinical neuroscience, 21(12):2065–2071, 2014.

5. Jan Kolčava, Jan Kočica, Monika Hulová, Ladislav Dušek, Magda Horáková, Miloš Kěrkovsky, Jakub Stulík, Marek Dostál, Matyas Kuhn, Eva Vlčková, et al. Conversion of clinically isolated syndrome to multiple sclerosis: a prospective study. Multiple Sclerosis and Related Disorders, 44:102262, 2020.

6. Fay Probert, Tianrong Yeo, Yifan Zhou, Megan Sealey, Siddharth Arora, Jacqueline Palace, Timothy DW Claridge, Rainer Hillenbrand, Johanna Oechtering, David Leppert, et al. Integrative biochemical, proteomics and metabolomics cerebrospinal fluid biomarkers predict clinical conversion to multiple sclerosis. Brain Communications, 3(2):fcab084, 2021.

7. Emmanuelle Waubant, Robyn Lucas, Ellen Mowry, Jennifer Graves, Tomas Olsson, Lars Alfredsson, and Annette Langer-Gould. Environmental and genetic risk factors for ms: an integrated review. Annals of clinical and translational neurology, 6(9):1905–1922, 2019.

8. Sara Collorone, Srikirti Kodali, and Ahmed T Toosy. The protective role of breastfeeding in multiple sclerosis: Latest evidence and practical considerations. Frontiers in Neurology, 13: 1090133, 2023.

9. Alan J Thompson, Brenda L Banwell, Frederik Barkhof, William M Carroll, Timothy Coetzee, Giancarlo Comi, Jorge Correale, Franz Fazekas, Massimo Filippi, Mark S Freedman, et al. Diagnosis of multiple sclerosis: 2017 revisions of the mcdonald criteria. The Lancet Neurology, 17(2):162–173, 2018.

10. G Bainaboina. Effects of multiple sclerosis on motor movement. J Mult Scler, 8(1), 2021.

11. Andrew J Solomon, Roman Pettigrew, Robert T Naismith, Salim Chahin, Stephen Krieger, and Brian Weinshenker. Challenges in multiple sclerosis diagnosis: Misunderstanding and misapplication of the mcdonald criteria. Multiple Sclerosis Journal, 27(2):250–258, 2021.

12. TK Banerjee, M Saha, E Ghosh, A Hazra, A Das, D Choudhury, S Ojha, A Haldar, A Mukherjee, SS Nandi, et al. Conversion of clinically isolated syndrome to multiple sclerosis: a prospective multi-center study in eastern india. Multiple Sclerosis Journal–Experimental, Translational and Clinical, 5(2):2055217319849721, 2019.

13. Ludwig Kappos, Mark S Freedman, Chris H Polman, Gilles Edan, Hans-Peter Hartung, David H Miller, Xavier Montalbán, Frederik Barkhof, Ernst-Wilhelm Radü, Lars Bauer, et al. Effect of early versus delayed interferon beta-1b treatment on disability after a first clinical event suggestive of multiple sclerosis: a 3-year follow-up analysis of the benefit study. The Lancet, 370(9585):389–397, 2007.

14. Mathias Due Buron, Thor Ameri Chalmer, Finn Sellebjerg, Ismael Barzinji, Danny Bech, Jeppe Romme Christensen, Mette Kirstine Christensen, Victoria Hansen, Zsolt Illes, Henrik Boye Jensen, et al. Initial high-efficacy disease-modifying therapy in multiple sclerosis: a nationwide cohort study. Neurology, 95(8):e1041–e1051, 2020.

15. Ben W Thrower. Clinically isolated syndromes: predicting and delaying multiple sclerosis. Neurology, 68(24_suppl_4):S12–S15, 2007.

16. Santosh Tirunagari, Senthilkumar Mohan, David Windridge, and Yashaswini Balla. Addressing challenges in healthcare big data analytics. In International Conference on Multi-disciplinary Trends in Artificial Intelligence, pages 757–765. Springer, 2023.

17. Massimo Filippi, Paolo Preziosa, Douglas L Arnold, Frederik Barkhof, Daniel M Harrison, Pietro Maggi, Caterina Mainero, Xavier Montalban, Elia Sechi, Brian G Weinshenker, et al. Present and future of the diagnostic work-up of multiple sclerosis: the imaging perspective. Journal of neurology, 270(3):1286–1299, 2023.

18. Vijay Simha Reddy Chennareddy, Santosh Tirunagari, Senthilkumar Mohan, David Windridge, and Yashaswini Balla. Interpretable chronic kidney disease risk prediction from clinical data using machine learning. In International Conference on Multi-disciplinary Trends in Artificial Intelligence, pages 683–691. Springer, 2023.

19. Haike Zhang, Esther Alberts, Viola Pongratz, Mark Mühlau, Claus Zimmer, Benedikt Wiestler, and Paul Eichinger. Predicting conversion from clinically isolated syndrome to multiple sclerosis–an imaging-based machine learning approach. NeuroImage: Clinical, 21:101593, 2019.

20. Youngjin Yoo, Lisa YW Tang, David KB Li, Luanne Metz, Shannon Kolind, Anthony L Traboulsee, and Roger C Tam. Deep learning of brain lesion patterns and user-defined clinical and mri features for predicting conversion to multiple sclerosis from clinically isolated syndrome. Computer Methods in Biomechanics and Biomedical Engineering: Imaging & Visualization, 7(3):250–259, 2019.

21. V Wottschel, DC Alexander, PP Kwok, DT Chard, MARIA LAURA Stromillo, NICOLA De Stefano, AJ Thompson, DH Miller, and O Ciccarelli. Predicting outcome in clinically isolated syndrome using machine learning. NeuroImage: Clinical, 7:281–287, 2015.

22. Saeid Rasouli, Mohammad Sedigh Dakkali, Reza Azarbad, Azim Ghazvini, Mahdi Asani, Zahra Mirzaasgari, and Mohammed Arish. Predicting the conversion from clinically isolated syndrome to multiple sclerosis: An explainable machine learning approach. Multiple Sclerosis and Related Disorders, 86:105614, 2024.

23. Yashaswini Balla, Santosh Tirunagari, and David Windridge. Pediatrics in artificial intelligence era: a systematic review on challenges, opportunities, and explainability. Indian Pediatrics, 60(7):561–569, 2023.

24. Víctor Chavarria, Guillermo Espinosa-Ramírez, Julio Sotelo, José Flores-Rivera, Omar Anguiano, Ana Campos Hernández, Edgar Daniel Guzmán-Ríos, Aleli Salazar, Graciela Ordoñez, and Benjamin Pineda. Conversion predictors of clinically isolated syndrome to multiple sclerosis in mexican patients: a prospective study. Archives of Medical Research, 54 (5):102843, 2023.

25. Renata Balnytė, Vaidas Matijošaitis, Ieva Čelpačenko, Miglė Malciūtė, Radvilė Stankevičiūtė, and Ovidijus Laucius. Factors related to the progression of clinically isolated syndrome to multiple sclerosis: A retrospective study in lithuania. Medicina, 58(9):1178, 2022.

26. Carl Kingsford and Steven L Salzberg. What are decision trees? Nature biotechnology, 26 (9):1011–1013, 2008.

27. Sandro Sperandei. Understanding logistic regression analysis. Biochemia medica, 24(1): 12–18, 2014.

28. Cha Zhang and Yunqian Ma. Ensemble machine learning, volume 144. Springer, 2012.

29. Claude Sammut and Geoffrey I Webb. Encyclopedia of machine learning. Springer Science & Business Media, 2011.

30. William S Noble. What is a support vector machine? Nature biotechnology, 24(12):1565–1567, 2006.

31. Tjalf Ziemssen, Katja Akgün, and Wolfgang Brück. Molecular biomarkers in multiple sclerosis. Journal of neuroinflammation, 16(1):272, 2019.

32. Karen K Chung, Daniel Altmann, Frederik Barkhof, Katherine Miszkiel, Peter A Brex, Jonathan O’Riordan, Michael Ebner, Ferran Prados, M Jorge Cardoso, Tom Vercauteren, et al. A 30-year clinical and magnetic resonance imaging observational study of multiple sclerosis and clinically isolated syndromes. Annals of neurology, 87(1):63–74, 2020.

33. Philip L De Jager, Lori B Chibnik, Jing Cui, Joachim Reischl, Stephan Lehr, K Claire Simon, Cristin Aubin, David Bauer, Jürgen F Heubach, Rupert Sandbrink, et al. Integration of genetic risk factors into a clinical algorithm for multiple sclerosis susceptibility: a weighted genetic risk score. The Lancet Neurology, 8(12):1111–1119, 2009.

34. Ruth Dobson, Sreeram Ramagopalan, and Gavin Giovannoni. The effect of gender in clinically isolated syndrome (cis): a meta-analysis. Multiple Sclerosis Journal, 18(5):600–604, 2012.

