## Supplementary Material: for "Interpretable Machine Learning for Predicting Multiple Sclerosis Conversion from Clinically Isolated Syndrome"

### Data Analysis

In the Lithuanian dataset, the age groups with the highest risk of developing MS are 35-40 years and 55-60 years. In the Mexican dataset, the age group with the highest risk of developing MS is 35-40 years, as shown in in Supplementary Material: Fig. S1.

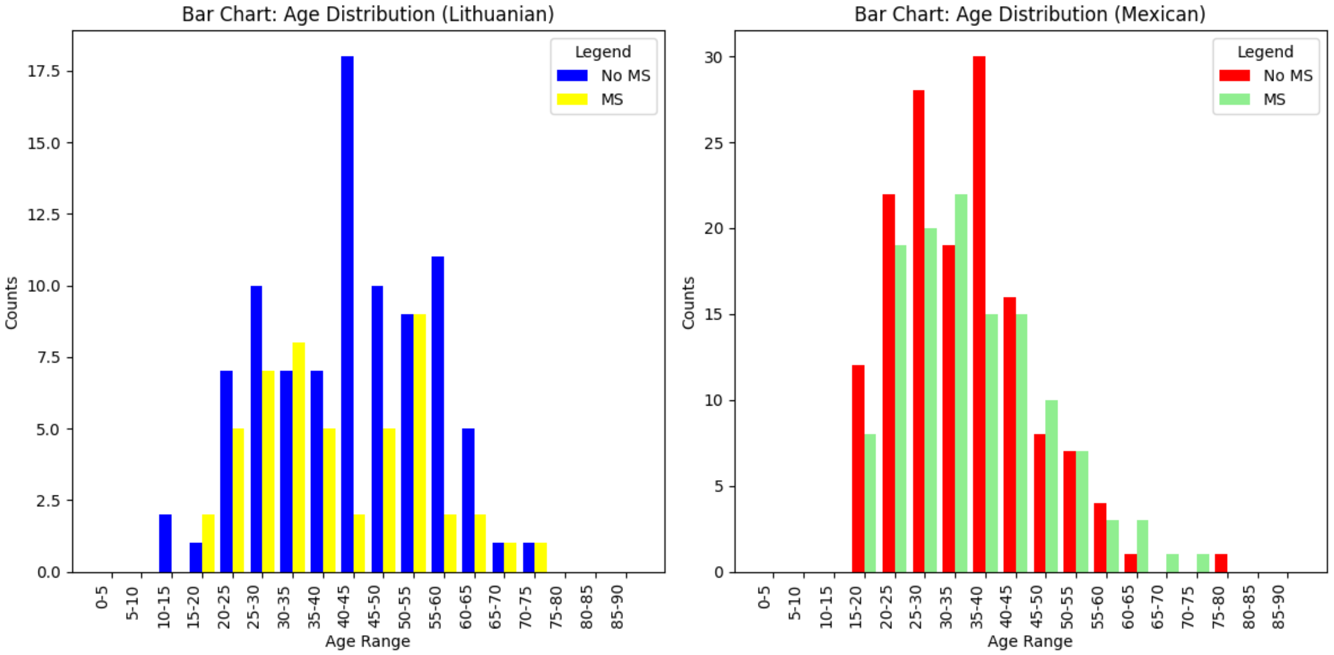


**Fig. S1.** Age-MS Distribution for Lithuanian (left) and Mexican (right) datasets.

In Lithuanian data, pathological reflexes, Babinski's sign, increased protein and IgG levels in CSF were multicollinear (Supplementary Material: Fig. S3). In the Mexican dataset, initial and final EDSS were multicollinear (Supplementary Material: Fig. S2).

In both the datasets, the number of healthy females outnumbered healthy males and the percentage of male MS patients exceeded female MS patients (Supplementary Material: Fig. S4). Oligoclonal bands in CSF significantly elevated MS risk (Supplementary Material: Fig. S5). Having MRI spinal lesions increased MS conversion risk (Supplementary Material: Fig. S6). VEP+ appeared more in MS patients (Supplementary Material: Fig. S7). In Lithuanian data, periventricular lesions did not significantly affect MS counts, while in the Mexican data they did (Supplementary Material: Fig. S8). MRI infratentorial lesions minimally impacted MS risk in the Lithuanian data but significantly raised MS risk in the Mexican data (Supplementary Material: Fig. S9). BAEP positivity was associated with higher MS risk (Supplementary Material: Fig. S10).

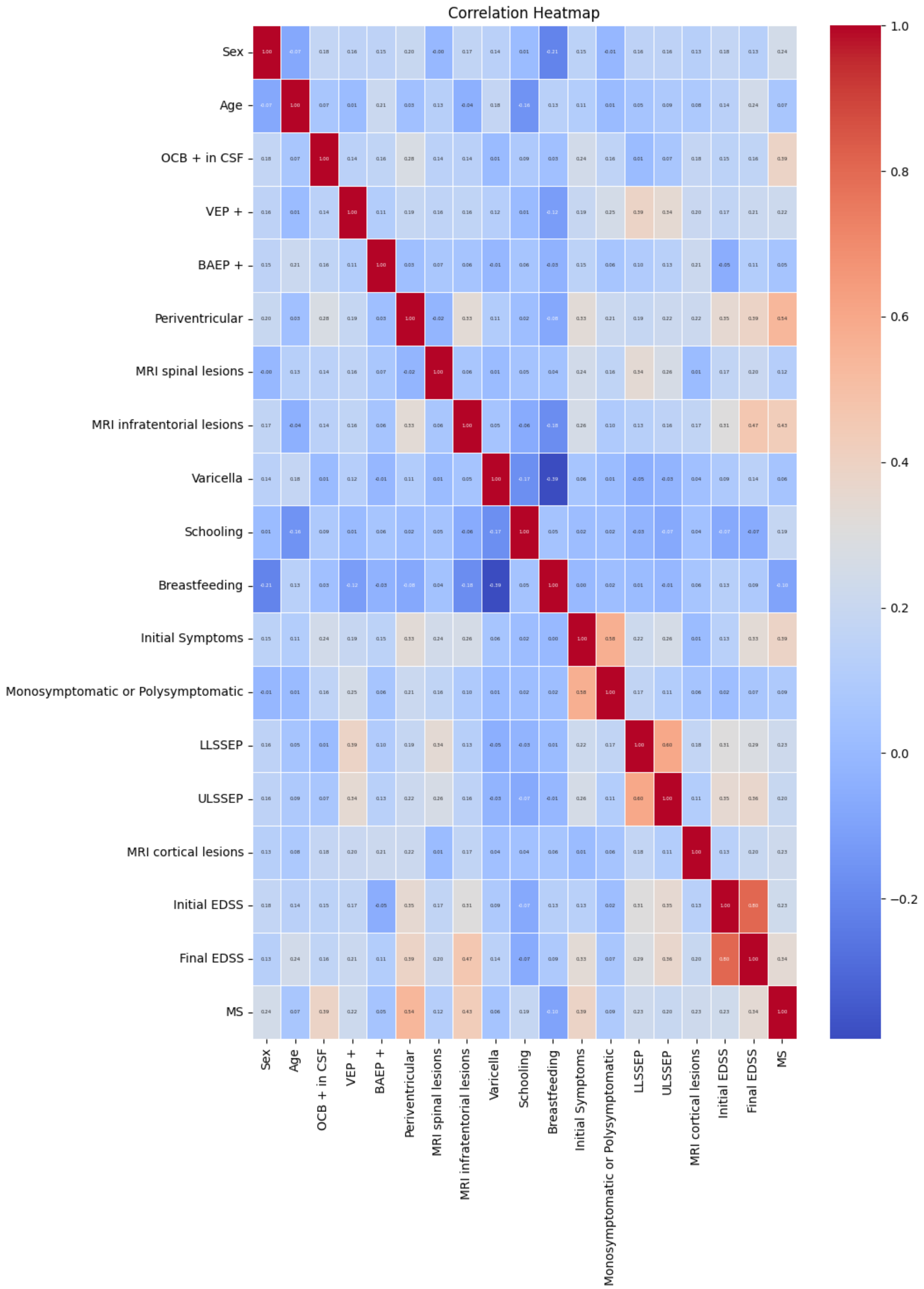


**Fig. S2.** Correlation Heatmap showing multicollinear features in the Mexican dataset.

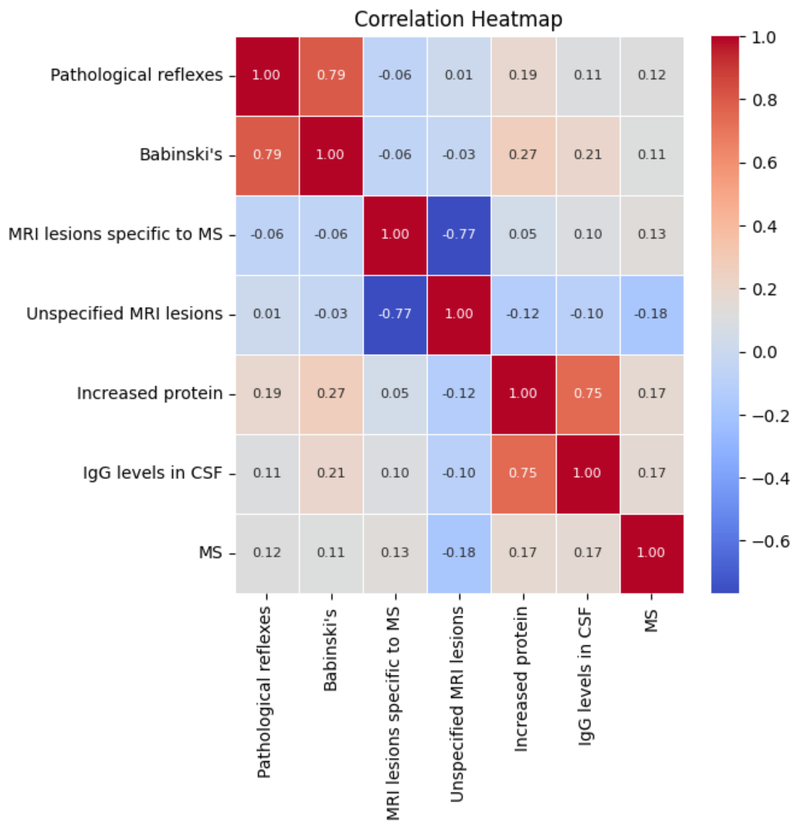


**Fig. S3.** Correlation Heatmap showing multicollinear features in the Lithuanian dataset.

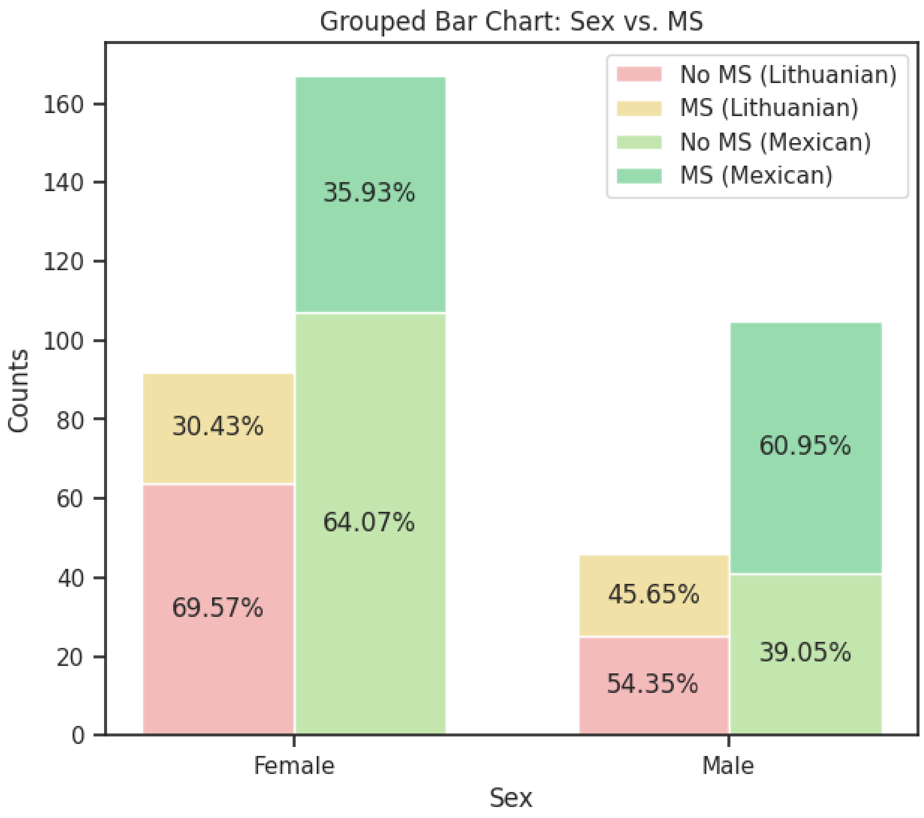


**Fig. S4.** Sex-MS Relationship for both datasets.

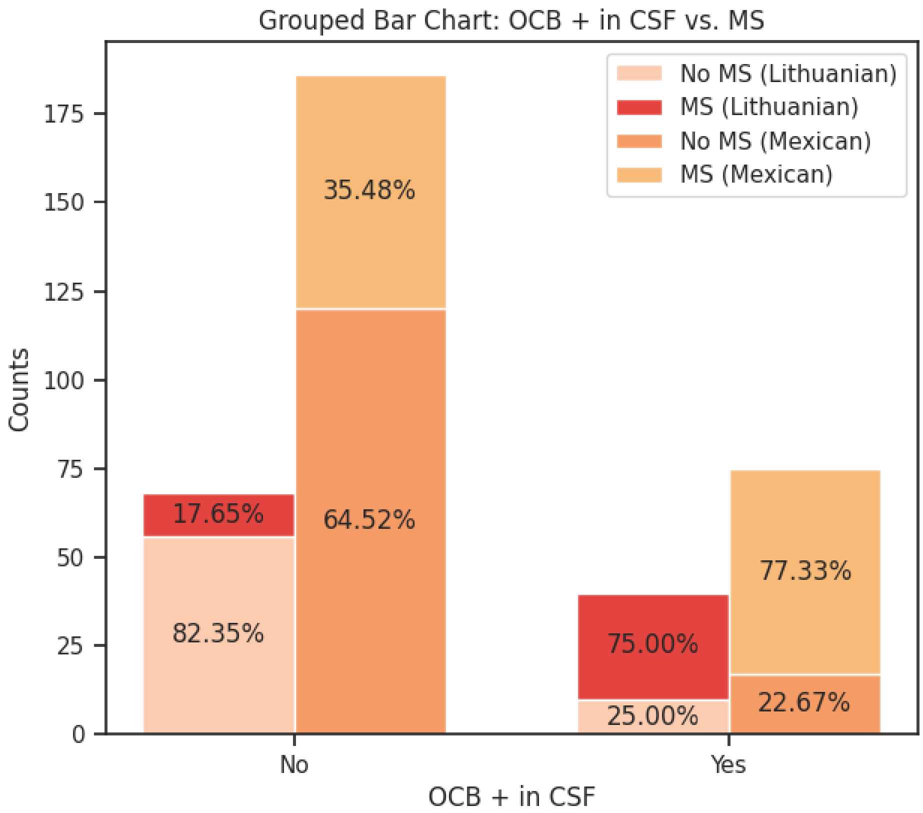


**Fig. S5.** OCB + in CSF-MS Relationship for both datasets.

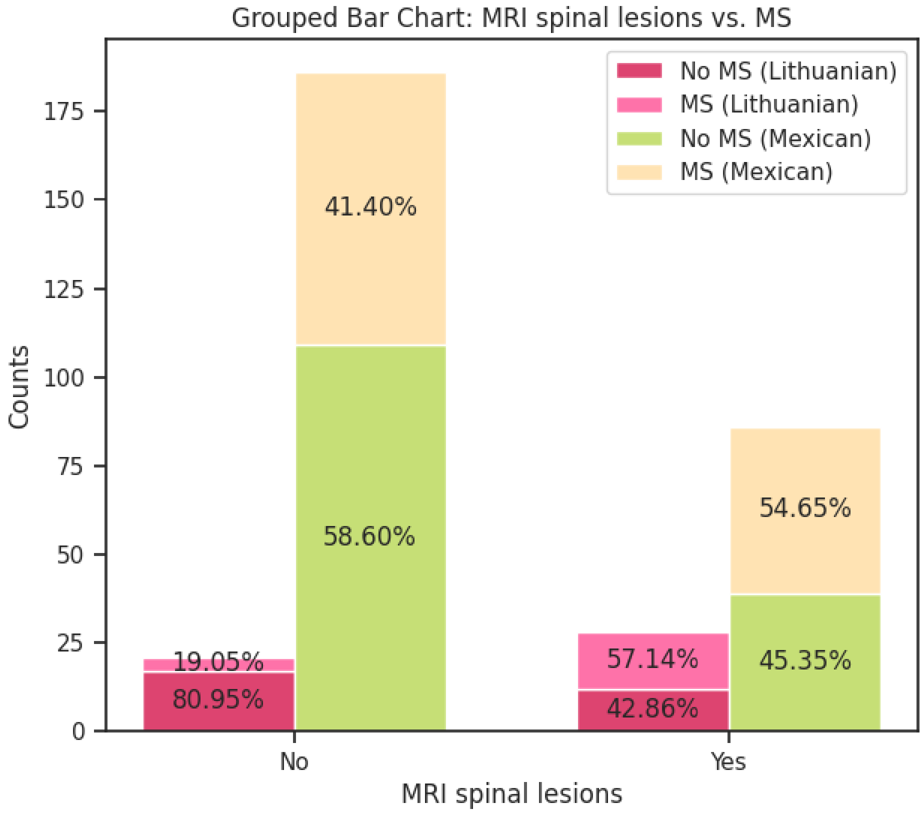


**Fig. S6.** MRI Spinal Lesions-MS Relationship for both datasets.

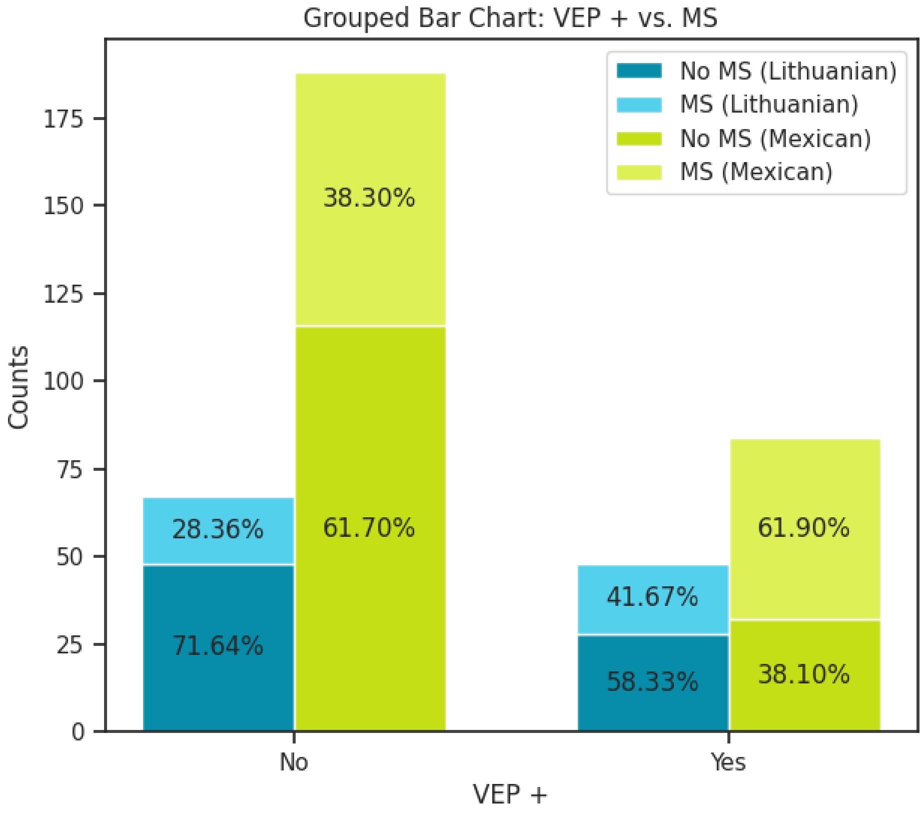


**Fig. S7.** VEP+ - MS Relationship for both datasets

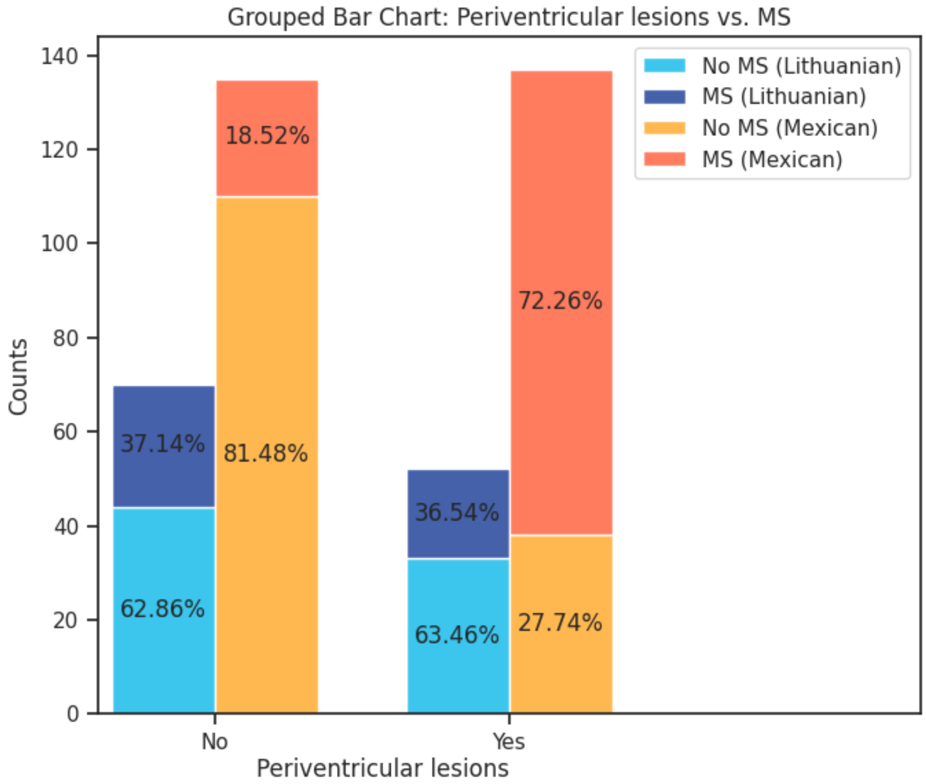


**Fig. S8.** Periventricular Lesions-MS Relationship for both datasets.

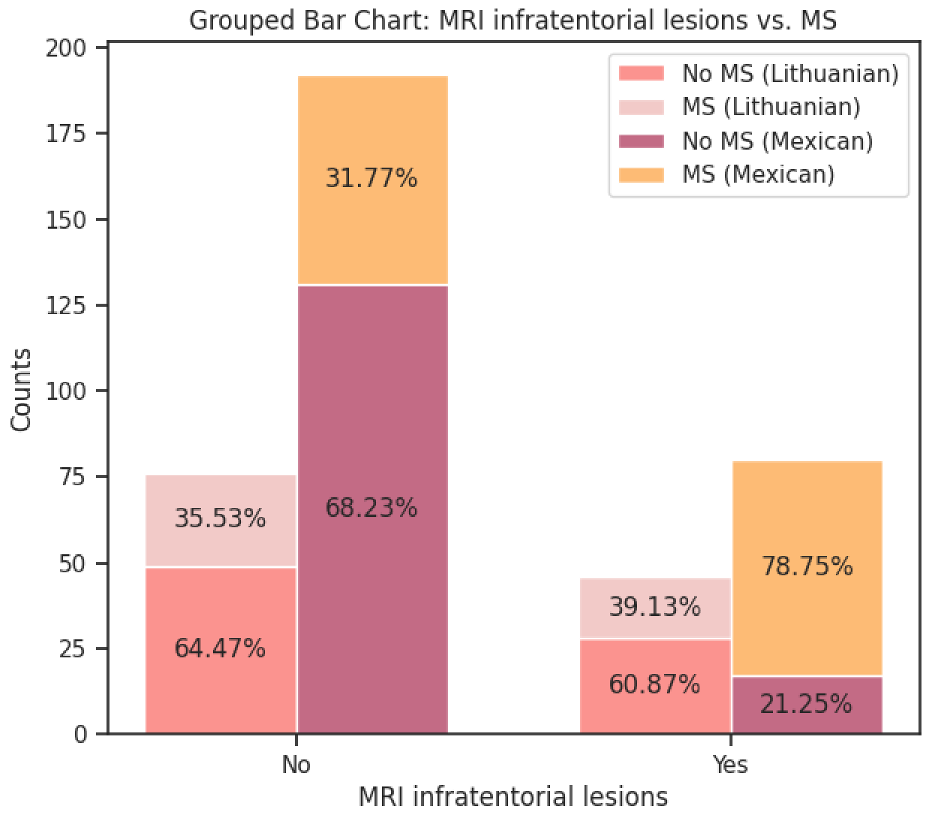


**Fig. S9.** MRI Infratentorial Lesions-MS Relationship for both datasets.

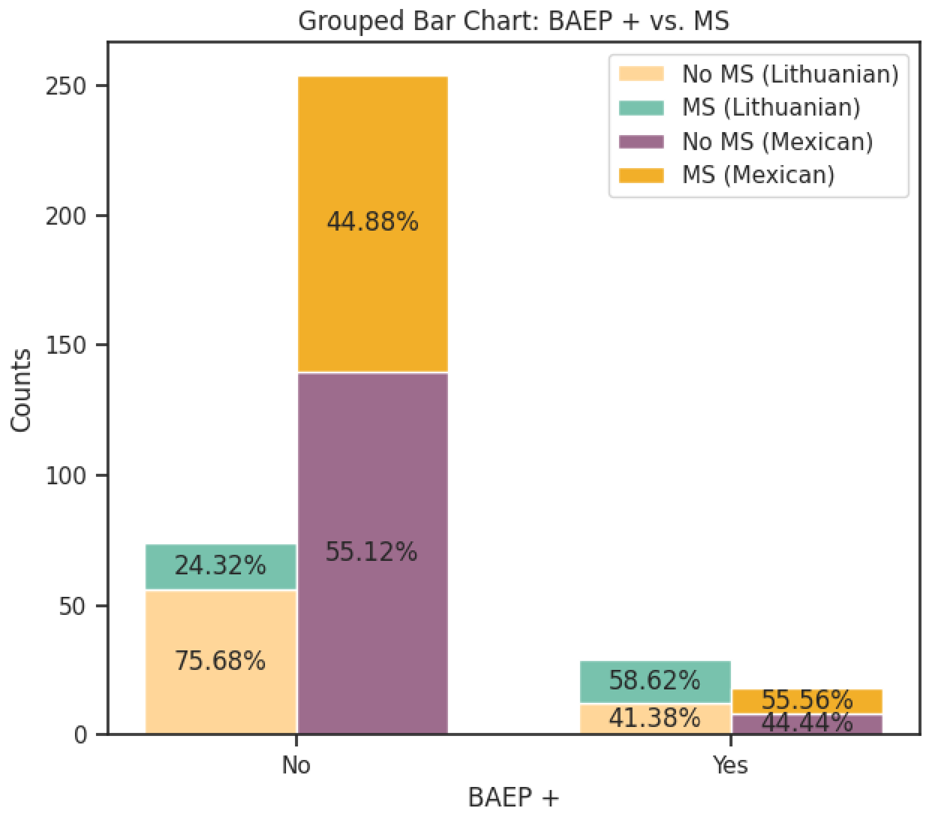


**Fig. S10.** BAEP +- MS Relationship for both datasets.

### Additional Results

| Imputation | Model | Data | F1 Score |
|---|---|---|---|
| **Simple Imputer** | NB | Train | 0.63 |
| | | Test | 0.47 |
| | DT | Train | 0.79 |
| | | Test | 0.79 |
| | RF | Train | 1.00 |
| | | Test | 0.71 |
| **EM Imputer** | NB | Train | 0.74 |
| | | Test | 0.79 |
| | DT | Train | 0.78 |
| | | Test | 0.79 |
| | RF | Train | 0.78 |
| | | Test | 0.79 |
| **MICE Imputer** | NB | Train | 0.74 |
| | | Test | 0.78 |
| | DT | Train | 0.83 |
| | | Test | 0.82 |
| | RF | Train | 0.94 |
| | | Test | 0.63 |

**Table S1.** F1 Scores of NB, DT and RF classifiers for Simple, EM and MICE Imputations.

| Imputation | PCA | Model | Data | F1 Score |
|---|---|---|---|---|
| Simple Imputer | Yes | NB | Train | 0.83 |
| | | | Test | 0.61 |
| | | DT | Train | 0.86 |
| | | | Test | 0.44 |
| | | RF | Train | 0.94 |
| | | | Test | 0.66 |
| EM Imputer | Yes | NB | Train | 0.84 |
| | | | Test | 0.62 |
| | | DT | Train | 0.61 |
| | | | Test | 0.50 |
| | | RF | Train | 0.90 |
| | | | Test | 0.57 |
| MICE Imputer | Yes | NB | Train | 0.81 |
| | | | Test | 0.78 |
| | | DT | Train | 0.65 |
| | | | Test | 0.50 |
| | | RF | Train | 0.93 |
| | | | Test | 0.56 |

**Table S2.** F1 Scores for all 3 models using Simple, EM and MICE Imputations with PCA.

### Ethical Approval

#### 8. Appendix: Research Ethics Approval Letter

Middlesex University London

Natural Science REC

The Burroughs
Hendon London NW4 4BT

Main Switchboard: 0208 411 5000

07/07/2023

**APPLICATION NUMBER:** 26123

Dear Eden Caroline Daniel and all collaborators/co-investigators

**Re: Your ethics application 26123**: Predicting Multiple Sclerosis Conversion from Clinically Isolated Syndrome using Machine Learning on Clinical Data

**Supervisor:** Santosh Tirunagari

**Co-investigators/collaborators**:

Thank you for submitting your application. I can confirm that your application has been given APPROVAL from the date of this letter by the Natural Science REC.

The following documents have been reviewed and approved as part of this research ethics application:

| Document Type | File Name | Date | Version |
|---|---|---|---|
| Data Access Approval | CC By 4.0 License-Mexican and Lithuanian Datasets | | CC By 4.0 |

Although your application has been approved, the reviewers of your application may have made some useful comments on your application. Please look at your online application again to check whether the reviewers have added any comments for you to look at.

Also, please note the following:

1.Please ensure that you contact your supervisor/research ethics committee (REC) if any changes are made to the research project which could affect your ethics approval. There is an Amendment sub-form on MORE that can be completed and submitted to your REC for further review.

2.You must notify your supervisor/REC if there is a breach in data protection management or any issues that arise that may lead to a health and safety concern or conflict of interests.

3.If you require more time to complete your research, i.e., beyond the date specified in your application, please complete the Extension sub-form on MORE and submit it your REC for review.

4.Please quote the application number in any correspondence.

5.It is important that you retain this document as evidence of research ethics approval, as it may be required for submission to external bodies (e.g., NHS, grant awarding bodies) or as part of your research report, dissemination (e.g., journal articles) and data management plan.

6.Also, please forward any other information that would be helpful in enhancing our application form and procedures - please contact to provide feedback.

Good luck with your research.

Yours sincerely,
Chair Natural Science REC

82

**Fig. S11.** Ethical approval for the use of datasets.
